# COVID-19 breakthrough infections and pre-infection neutralizing antibody

**DOI:** 10.1101/2021.10.20.21265301

**Authors:** Shohei Yamamoto, Kenji Maeda, Kouki Matsuda, Akihito Tanaka, Kumi Horii, Kaori Okudera, Junko S. Takeuchi, Tetsuya Mizoue, Maki Konishi, Mitsuru Ozeki, Haruhito Sugiyama, Nobuyoshi Aoyanagi, Hiroaki Mitsuya, Wataru Sugiura, Norio Ohmagari

**Author notes:** **Address for the corresponding author:** Tetsuya Mizoue, MD, PhD, Department of Epidemiology and Prevention, National Center for Global Health and Medicine, 1-21-1, Toyama, Shinjuku-ku, Tokyo, 162-8655, Japan.

## Abstract

**Background:** While increasing coverage of effective vaccines against coronavirus disease 2019 (COVID-19), emergent variants raise concerns about breakthrough infections. Data are limited, however, whether breakthrough infection during the epidemic of the variant is ascribed to insufficient vaccine-induced immunogenicity.

**Methods:** We described incident COVID-19 in relation to the vaccination program among workers of a referral hospital in Tokyo. During the predominantly Delta epidemic, we followed 2,473 fully vaccinated staff (BNT162b2) for breakthrough infection and selected three matched controls. We measured pre-infection neutralizing antibodies against the wild-type, Alpha (B.1.1.7), and Delta (B.1.617.2) strains using live viruses and anti-spike antibodies using quantitative assays, and compared them using the generalized estimating equation model between the two groups.

**Results:** No COVID-19 cases occurred 1–2 months after the vaccination program during the fourth epidemic wave in Japan, dominated by the Alpha variant, while 22 cases emerged 2–4 months after the vaccination program during the fifth wave, dominated by the Delta variant. In the vaccinated cohort, all 17 cases of breakthrough infection were mild or asymptomatic and had returned to work early. There was no measurable difference between cases and controls in pre-infection neutralizing antibody titers against the wild-type, Alpha, and Delta, and anti-spike antibody titers, while neutralizing titers against the variants were considerably lower than those against the wild-type.

**Conclusions:** Pre-infection neutralizing antibody titers were not decreased among patients with breakthrough infection under the Delta variant rampage. The result points to the importance of infection control measures in the post-vaccination era, irrespective of immunogenicity profile.

## Introduction

Clinical trials show that the mRNA-based vaccines against severe acute respiratory syndrome coronavirus 2 (SARS-CoV-2) are highly effective in lowering the risk of coronavirus disease (COVID-19).^1,2^ In countries with high coverage of vaccination programs, however, the number of COVID-19 patients has started growing, which has been attributed to the emergence of Variants of Concern, especially the Delta (B.1.617.2) variant,^3,4^ and the waning of vaccine efficacy over time.^5,6^

Epidemiological evidence is scarce regarding the role of vaccine-induced immunogenicity against Variants of Concern. In a case-control study among vaccinated healthcare workers, patients with breakthrough infection during the epidemic of the Alpha (B.1.1.7) variant had significantly lower peri-infection neutralizing antibody titers than controls.^7^ Similar result was reported from a Vietnamese study^8^ on breakthrough infection during the epidemic of the Delta variant, which is more resistant to the current vaccines than the Alpha variant.^9^

In Japan, the vaccination program has started in mid-February 2021, initially among healthcare workers using the BNT162b2 mRNA vaccine (Pfizer-BioNTech). After that, Japan has been hit by the two large waves of the COVID-19 epidemic: the fourth wave (April and May 2021), dominated by the Alpha variant, and the fifth wave (from July to September), overwhelmed with the Delta variant.^10^ This has prompted us to describe COVID-19 incidence in relation to the timing of the vaccination program and local epidemic wave among the staff of a large referral hospital in Tokyo, Japan, and compare pre-infection neutralizing antibodies against the wild-type, Alpha, and Delta strains between breakthrough infection cases and their matched controls during the large epidemic of the Delta variant.

## Methods

### Study setting and population

National Center for Global Health and Medicine (NCGM), comprised of two hospitals and affiliated facilities, is a medical research center for specific areas, including infectious disease. As their mission, NCGM has played a major role in the care and research of COVID-19 since the early stage of the epidemic^11^ and has accepted many severe COVD-19 patients. During the in-house vaccination program using COVID-19 mRNA-LNP BNT162b2 (Pfizer-BioNTech) from March through June 2021, more than 90% of the NCGM staff have received the two-dose vaccine.

In NCGM, a longitudinal serological study has been launched since July 2020 to monitor the spread of SARS-CoV-2 infection among the staff. The results of the first and second surveys have been reported elsewhere.^12,13^ In the third survey in June 2021 (within 4 months following the in-house vaccination program), we measured anti-SARS-CoV-2 spike protein antibodies, stored serum samples at -80 □ for further investigation, and collected data on COVID-19 (vaccination, occupational risk of SARS-CoV-2 infection, and infection prevention behaviors, etc.) via a questionnaire. Of the 3,072 workers invited, 2,779 (90%) participated. Written informed consent was obtained from all participants, and the study procedure was approved by the NCGM ethics committee (the approved number: NCGM-G-003598).

### Identification of COVID-19 cases

We identified COVID-19 cases among the study participants against the COVID-19 patients records kept by the NCGM Hospital Infection Prevention and Control Unit, which provided information on data of diagnosis, diagnostic procedure, possible route of the infection (close contact), symptoms, hospitalization, and return to work for all cases; and virus strain and Ct values for those who were diagnosed at NCGM. For COVID-19 cases among non-participants, only the date of diagnosis was informed anonymously.

### Case-control selection

We conducted a case-control matching analysis within a sub-cohort of the participants of the third serological survey. Participants to be included must have completed two doses of the vaccination before the survey (n=2,456), while excluded if those who had a history of COVID-19 (n=14), showed positive anti-SARS-CoV-2 antibody test at any serological survey (n=40), leaving 2,415 in the sub-cohort. Breakthrough infection was defined if COVID-19 was diagnosed at least 14 days after the second dose of vaccine, and the serum sample of those identified through September 6, 2021 was measured. To ensure the statistical power of the study, we applied a case-control ratio of 1:3. For each case, we selected three un-infected controls from the sub-cohort while matching worksite, sex, age, the interval between the second vaccination and blood sampling, and the propensity score, which was created based on body mass index, occupational exposure risk of SARS-CoV-2, and several infection prevention/risk behaviors. The details of the case-control matching algorithm are described in **Supplemental Text 1**.

### Neutralizing Antibody testing

The neutralizing activity of serum of cases and selected controls was determined by quantifying the serum-mediated suppression of the cytopathic effect (CPE) of each SARS-CoV-2 strain in VeroE6_TMPRSS2_ cells.^14^ The obtaining routes of the cells and each virus are described in **Supplemental Text 2**. Each serum was 4-fold serially diluted in a culture medium. The diluted sera were incubated with 100 50% tissue culture infectious dose (TCID_50_) of viruses at 37°C for 20 min (final serum dilution range of 1:20 to 1:4000), after which the serum-virus mixtures were inoculated to VeroE6_TMPRSS2_ cells (1.0 × 10^4^/well) in 96-well plates. For SARS-CoV-2 strains used in these assays are as follows: a Wuhan, wild-type strain (SARS-CoV-2_05-2N_, PANGO lineage B),^14^ an Alpha variant (SARS-CoV-2_QHN001_), and a Delta variant (SARS-CoV-2_1734_). After culturing the cells for three days, the levels of CPE observed in SARS-CoV-2-exposed cells were determined using the WST-8 assay employing Cell Counting Kit-8 (Dojindo, Kumamoto, Japan). The serum dilution that gave 50% inhibition of CPE was defined as the 50% neutralization titer (NT_50_). Each serum was tested in duplicate. The assayers were blinded to which serum was cases or controls.

### SARS-CoV-2 antibody testing

We assessed anti-SARS-CoV-2 antibodies for all participants of the third survey and retrieved those data for the case-control subsets. We quantitatively measured antibodies against the receptor-binding domain (RBD) of the SARS-CoV-2 spike protein by using the AdviseDx SARS-CoV-2 IgG II assay (Abbott) (IgG) and Elecsys Anti-SARS-CoV-2 S RUO (Roche) (predominantly IgG, also IgA and IgM). We also qualitatively measured antibodies against SARS-CoV-2 nucleocapsid protein using the SARS-CoV-2 IgG assay (Abbott) and Elecsys Anti-SARS-CoV-2 RUO (Roche), and used these data to exclude those with possible infection in the past.

### Statistical analysis

We compared the participants’ characteristics between cases and matched controls using Mann–Whitney U test or Fisher’s exact test as appreciate. To examine the antibody-mediated immune response at the pre-breakthrough infection, we compared their log-transformed titers of neutralizing (wild-type, Alpha, and Delta) and anti-spike antibodies using a generalized estimating equation (GEE) with the group assignment (case or control) and robust variance estimator. Then, we back-transformed and presented these values in geometric mean titer (GMT) with 95% confidence intervals (CIs). To compare the inter-individual differences in neutralizing antibody titers against the wild-type, Alpha, and Delta strains, we used the Wilcoxon ranked-sign test with adjustment using the Bonferroni method for multiple tests. Statistical analysis was performed using Stata 17.0 (StataCorp LLC), and graphics were made by GraphPad Prism 9 (GraphPad Inc). All *P* values were 2-sided, and *P* < 0.05 was considered statistically significant.

## Results

### Description of COVID-19 incidence (Figure 1)

**Figure 1.**
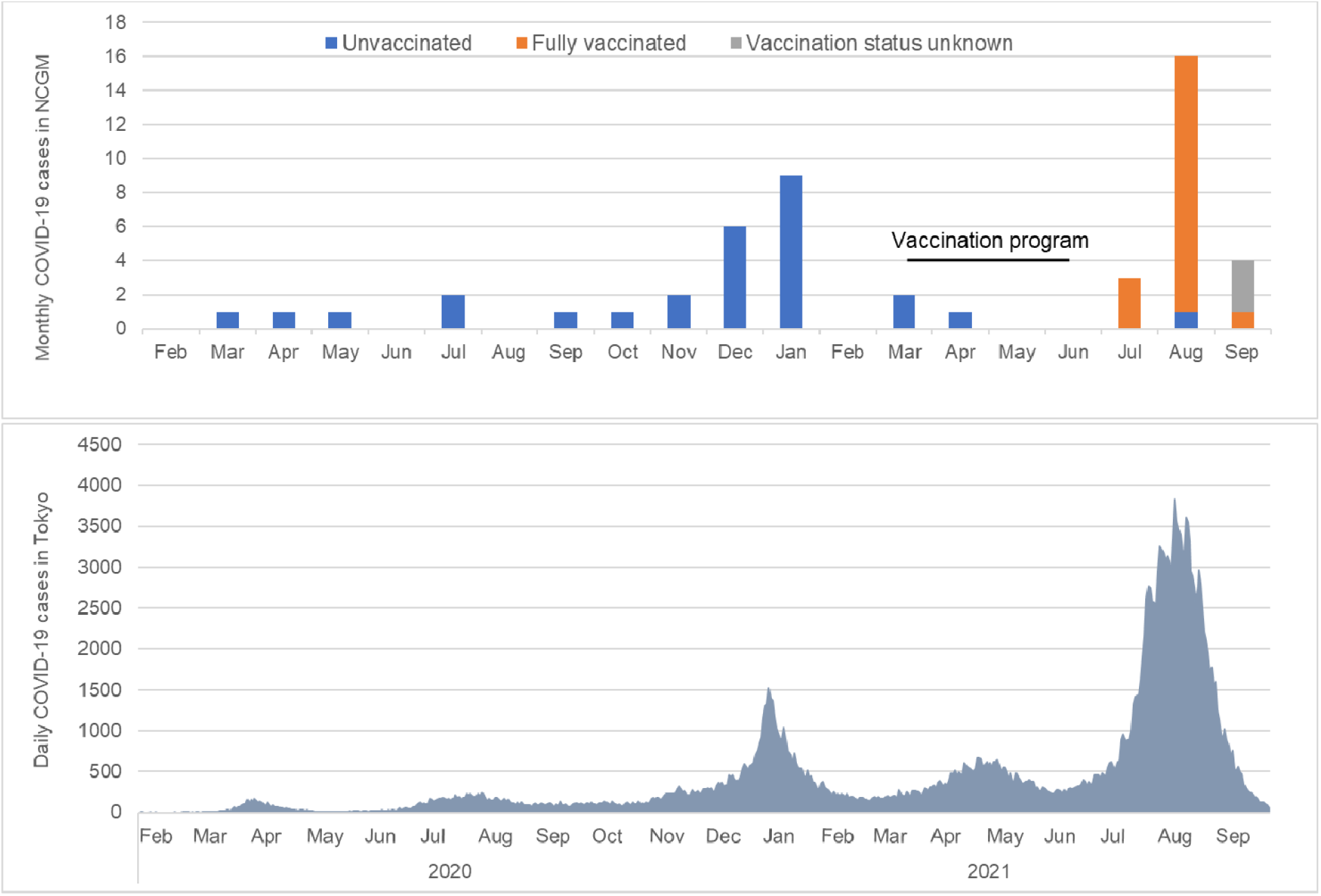
The number of confirmed COVID-19 cases at the National Center for Global Health (NCGM) and Tokyo between February 2020 and September 2021. The upper panel indicates the monthly confirmed COVID-19 cases among the NCGM workers. The lower panel indicates the daily confirmed COVID-19 cases in Tokyo, Japan, where the NCGM is located. In the NCGM, the vaccination program was done in March-April 2021 in Toyama (wherein 80% of NCGM staff) ward and April-Jun 2021 in Kohnodai ward (20% of NCGM staff).

Before the in-hospital vaccination program, 17 cases of COVID-19 (5.9 per 1000 persons) occurred among NCGM staff between November 2020 and February 2021 during the third wave of epidemic in Japan, dominated by the Japan-specific B.1.1.214 variant. During Japan’s fourth wave between April and May 2021 (within 2 months after the vaccination program in the Toyama ward [80% of NCGM staff]), no cases were identified among staff in that ward. During the fifth epidemic wave between July and September 2021, when the Delta was the dominant strain, 22 cases were identified (7.5 per 1000 persons). Of 19 cases who agreed to provide information about vaccination status, 18 (95%) were fully vaccinated. The risk of infection during the two months in the fifth wave (after the hospital vaccination program) was only slightly higher than that observed during the third wave (December 2020 to January 2021) (before vaccination) (5.2 per 1000 persons), while the number of cases in Tokyo during the fifth wave (n=133,989) was 3.5 times higher than that during the third wave (n=38,492).

### Characteristics of breakthrough infection in a case-control analysis

Of 17 cases of breakthrough infection in the case-control analysis, 47% were men, the median age was 29 (interquartile range [IQR]: 25–44) years, and the median interval between the second COVID-19 vaccination and breakthrough infection was 111 (IQR: 98–123) days. The major occupations included nurses (53%), allied healthcare professionals (24%), and doctors (18%), and more than half were at low occupational risk of SARS-CoV-2 infection (59%). The majority (88–100%) showed good adherence to infection prevention practices. For 41% of cases, they commute to work by public transportation five or more days per week. As regards risky behaviors on leisure, 18% reported that they had experienced spending more than 30 minutes in the 3Cs (crowded places, close-contact settings, and confined and enclosed spaces) without wearing a mask, and 12% reported eaten with five or more people that lasted at least an hour. There was no significant difference in these figures between cases and controls (**Table 1**).

**Table 1.**
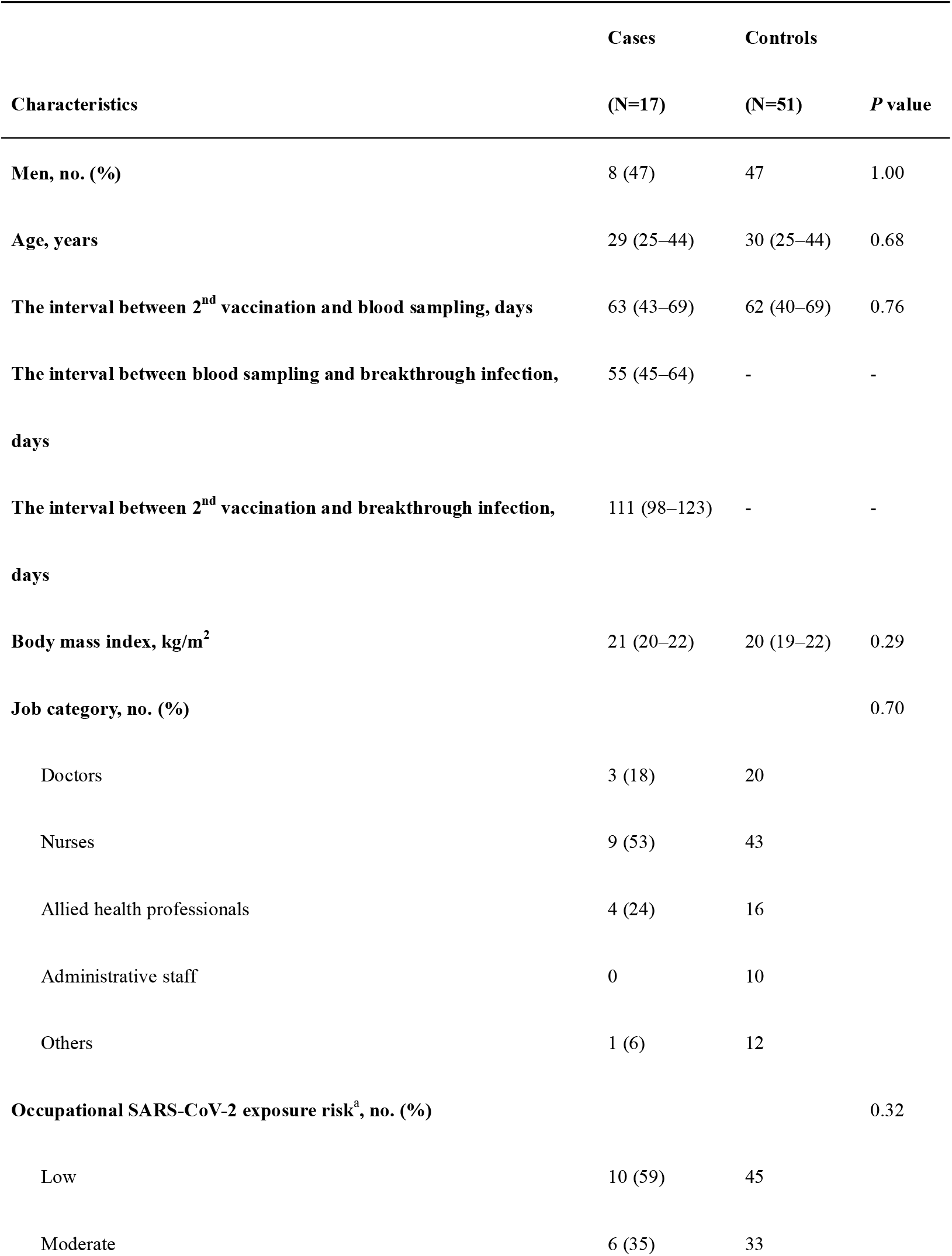
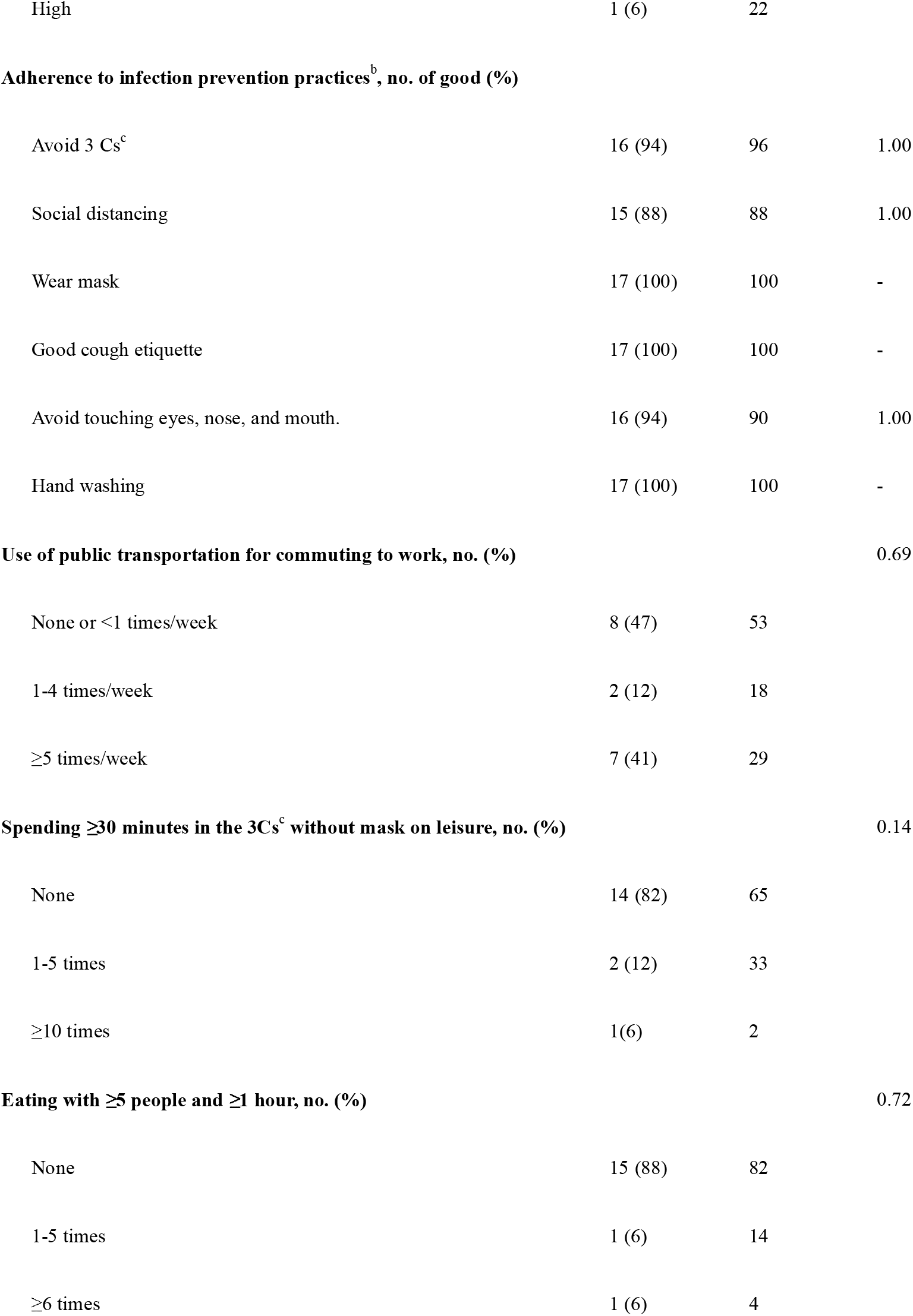
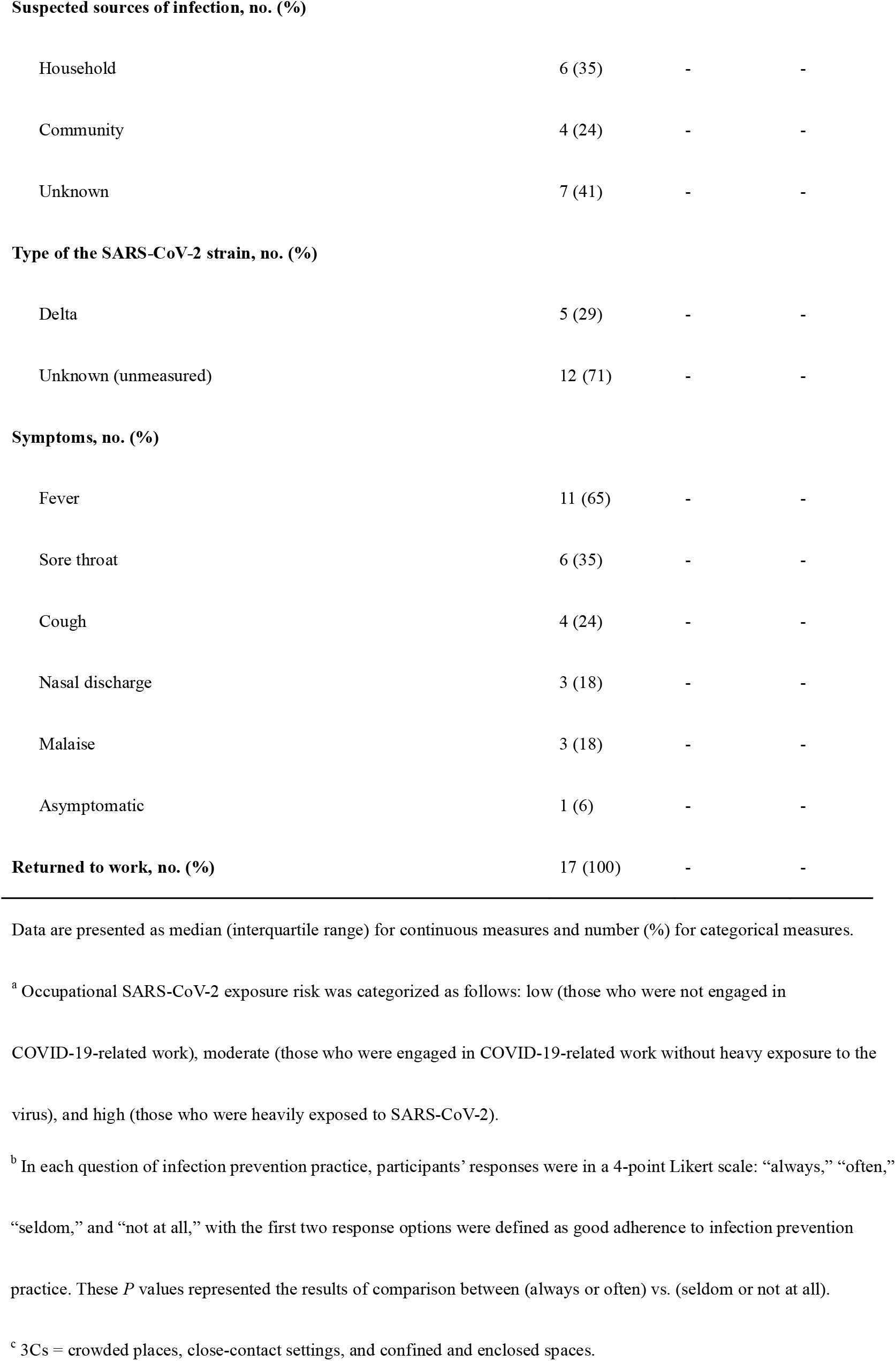
Characteristics of participants in the case-control study

All the breakthrough cases occurred independently in different timings and departments, and there was no evidence of the clustering of infection in the hospital. The suspected sources of breakthrough infection were contacting COVID-19 patients in the household (35%), those in the community (24%), and unknown (41%), while none were suspected of nosocomial infection. Data on the type of SARS-CoV-2 strain is available for only 5 cases, and all of those were the Delta variant. Their symptom was as follows: fever (65%); sore throat (35%); cough (24%); nasal discharge (18%); malaise (18%). All cases (except one asymptomatic case) had only mild symptoms and have returned to work without hospitalization or special medical care (**Table1**). The detailed characteristics of each breakthrough infection case are described in **Supplemental Table 1**.

### Neutralizing and anti-spike antibodies

The pre-infection neutralizing antibody titers against the wild-type, Alpha, and Delta (median 62 days between the second vaccination and blood sampling) were not statistically different between breakthrough cases and their matched controls (**Table 2 and Figure 2A-C**). The GEE predicted neutralizing antibody titers against the wild-type was 405 (95% confidence interval [CI], 327-501) for cases and 408 (95% CI, 320–520) for controls, and this predicted case-to-control ratio was 0.99 (95% CI, 0.74–1.34). Those against the Alpha variant were 116 (95% CI, 80–169) for cases and 122 (95% CI, 96–155) for controls, with the ratio of 0.95 (95%CI, 0.71–1.28). Those against the Delta variant were 123 (95% CI, 85–177) for cases and 135 (95% CI, 108–170) for controls, and the ratio was 0.91 (95% CI, 0.61–1.34).

**Table 2.**
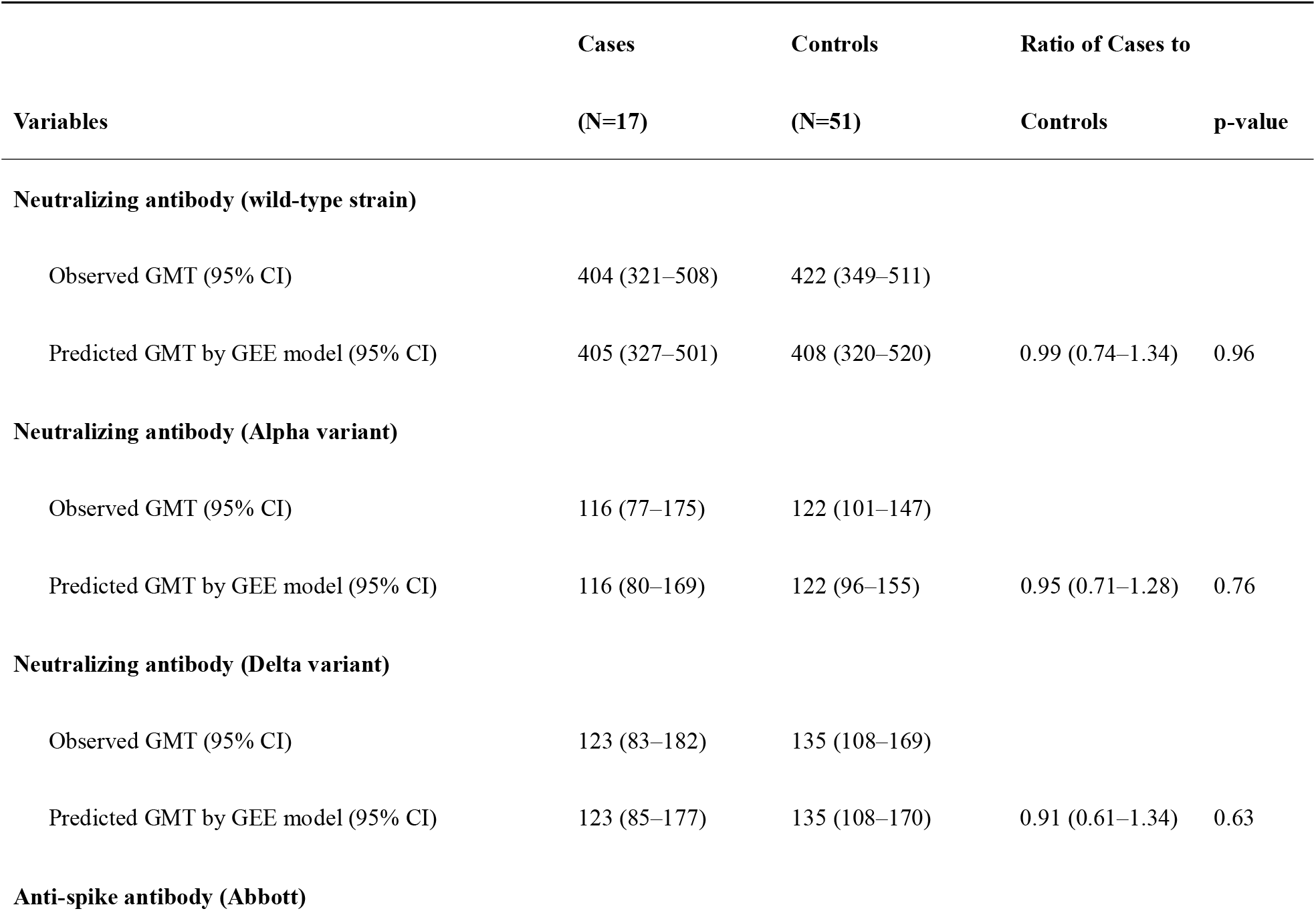
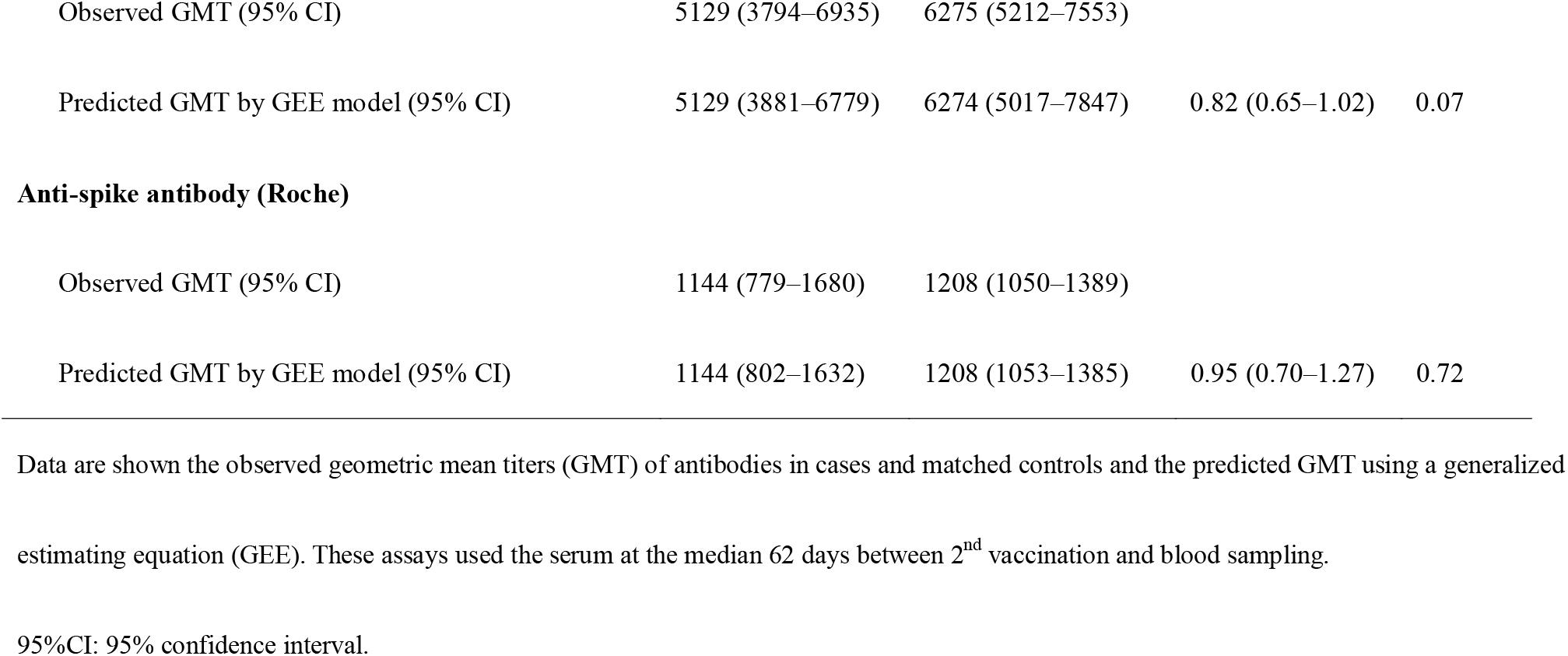
Comparison of pre-infection antibody titers between cases and controls

| Variables | Cases | Controls | Ratio of Cases to |  |
| --- | --- | --- | --- | --- |
|  | (N=17) | (N=51) | Controls | p-value |
| **Neutralizing antibody (wild-type strain)** |  |  |  |  |
| Observed GMT (95% CI) | 404 (321–508) | 422 (349–511) |  |  |
| Predicted GMT by GEE model (95% CI) | 405 (327–501) | 408 (320–520) | 0.99 (0.74–1.34) | 0.96 |
| **Neutralizing antibody (Alpha variant)** |  |  |  |  |
| Observed GMT (95% CI) | 116 (77–175) | 122 (101–147) |  |  |
| Predicted GMT by GEE model (95% CI) | 116 (80–169) | 122 (96–155) | 0.95 (0.71–1.28) | 0.76 |
| **Neutralizing antibody (Delta variant)** |  |  |  |  |
| Observed GMT (95% CI) | 123 (83–182) | 135 (108–169) |  |  |
| Predicted GMT by GEE model (95% CI) | 123 (85–177) | 135 (108–170) | 0.91 (0.61–1.34) | 0.63 |
| **Anti-spike antibody (Abbott)** |  |  |  |  |
| Observed GMT (95% CI) | 5129 (3794–6935) | 6275 (5212–7553) |  |  |
| Predicted GMT by GEE model (95% CI) | 5129 (3881–6779) | 6274 (5017–7847) | 0.82 (0.65–1.02) | 0.07 |
| **Anti-spike antibody (Roche)** |  |  |  |  |
| Observed GMT (95% CI) | 1144 (779–1680) | 1208 (1050–1389) |  |  |
| Predicted GMT by GEE model (95% CI) | 1144 (802–1632) | 1208 (1053–1385) | 0.95 (0.70–1.27) | 0.72 |

Data are shown the observed geometric mean titers (GMT) of antibodies in cases and matched controls and the predicted GMT using a generalized
estimating equation (GEE). These assays used the serum at the median 62 days between 2<sup>nd</sup> vaccination and blood sampling.
95%CI: 95% confidence interval.

**Figure 2.**
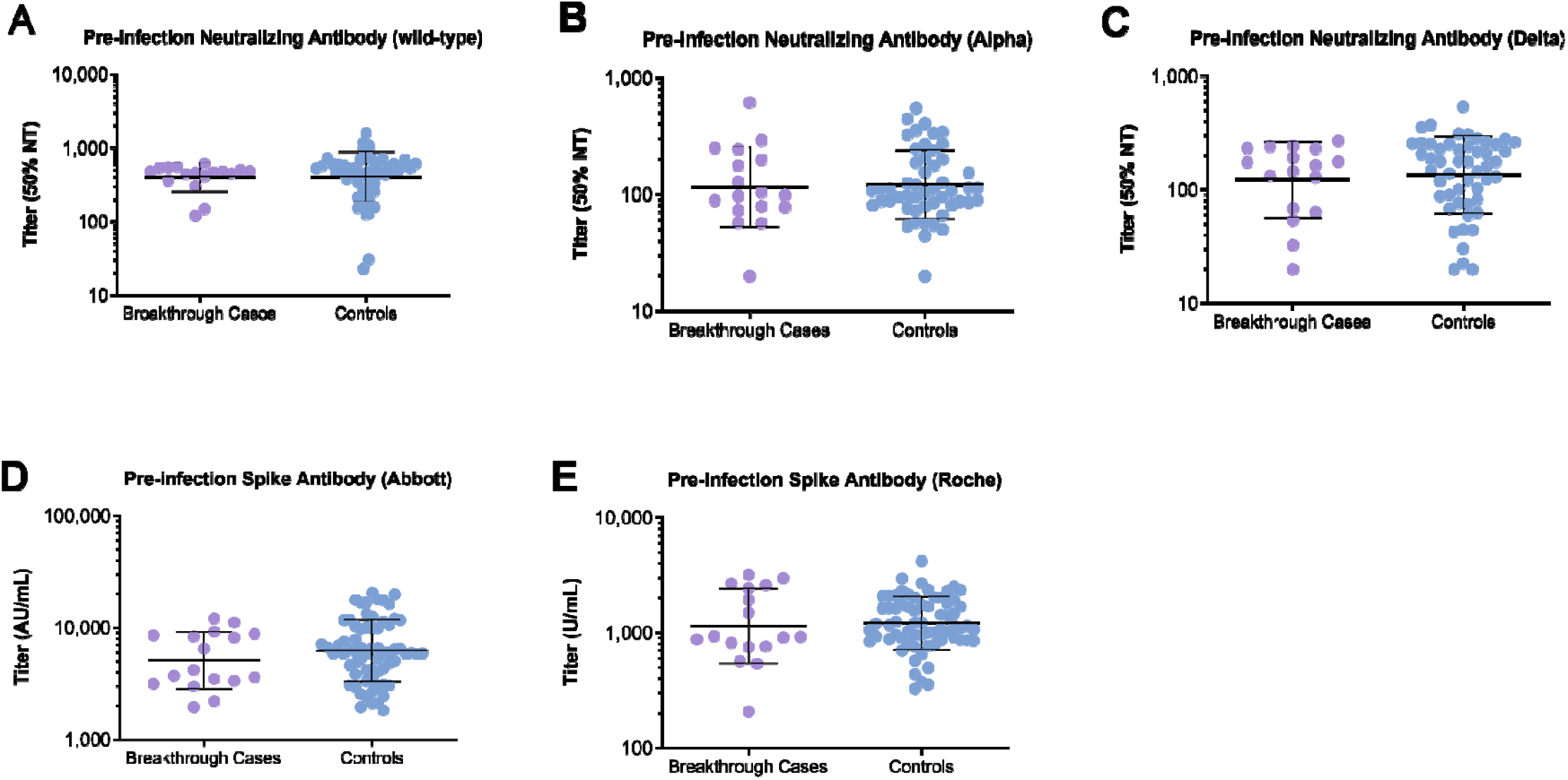
Pre-infection neutralizing and anti-spike antibody titers among cases and controls. Among the 17 fully vaccinated healthcare workers who had breakthrough infection with SARS-CoV-2 and the 51 matched controls, shown are neutralizing antibody titers against the wild-type strain (Panel A), the Alpha variant (Panel B), and the Delta variant (Panel C) during the pre-infection period (median 62 days since the second vaccination). Besides, shown were the comparison of anti-spike antibody titers measured by Abbott reagent (Panel D) and those by Roche reagent (Panel E) in the two groups. Each case of breakthrough infection was matched with three controls according to the worksite, sex, age, the interval between the second vaccination and blood sampling, and propensity score, estimated by body mass index, occupational exposure risk of SARS-CoV-2, and adherence to several infection preventive/risky behaviors. In each panel, the horizontal bars indicate the geometric means titers, and I bars indicate geometric standard deviations.

The predicted GMT of pre-infection anti-spike antibody titers were comparable among cases and controls (**Table 2 and Figure 2D-E**). The predicted GMT of pre-infection anti-spike antibody on the Abbott assay was 5129 (95% CI, 3881-6779) for cases and 6274 (95% CI, 5017-7847) for controls, for a ratio of 0.82 (95% CI, 0.65–1.02). That on the Roche assay was 1144 (95% CI, 802-1632) for cases and 1208 (95% CI, 1053-1385) for controls, and the ratio was 0.95 (95% CI, 0.70–1.27).

The neutralization titers against the Alpha and Delta variants were much lower than those against the wild-type (**Figure 3**). Among cases, the GMT (95% CI) of neutralizing antibody was 116 (77-175), 123 (83-182), and 404 (321-508) for the Alpha and Delta variants and the wild-type, respectively (*P* <0.01 for each variant versus Wild-type). Similar results were obtained for controls.

**Figure 3.**
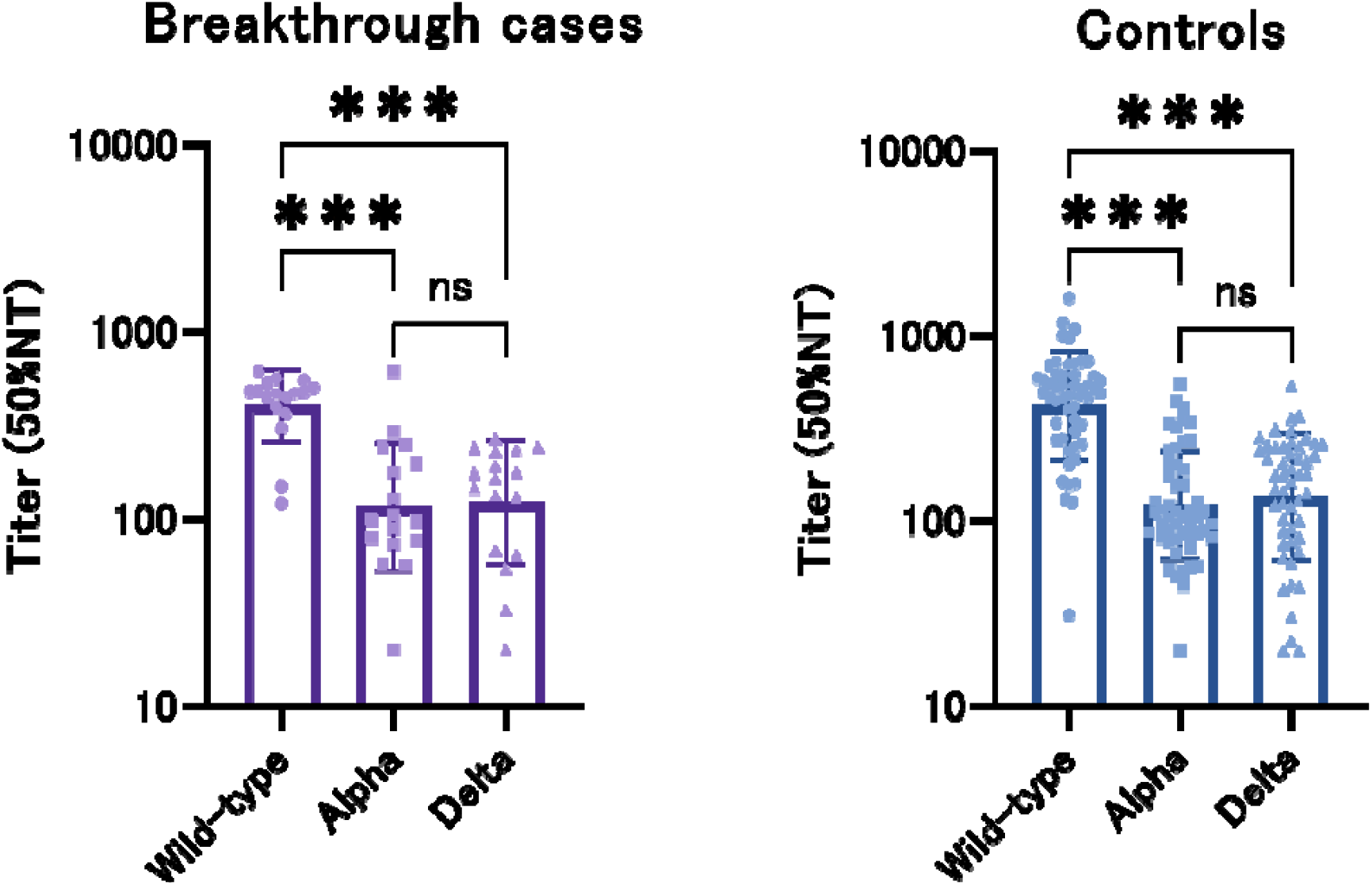
Neutralization of Delta (B.1.617.2) and Alpha (B.1.1.7) live viruses by serum from breakthrough cases (n=17) or controls (n=51) in comparison with the wild-type (Wuhan strain). Shown are the 50% neutralizing titer (50%NT), the serum dilution required for 50% virus inhibition, expressed as the geometric mean titers (GMT) with geometric standard deviations (I bars). The purple bars indicate breakthrough infections (n=17), and the blue bars indicate the matched controls (n=51). ***: p<0.001 by the Wilcoxon matched-pairs signed-rank test, ns: not significant.

## Discussion

Among the staff of a large referral hospital in Tokyo, a series of breakthrough infection has been documented during the largest COVID-19 epidemic wave, dominated by the Delta variant. In a case-control study nested within the cohort of vaccinated staff, pre-infection neutralizing antibody titers did not materially differ between breakthrough infection cases and their matched controls.

In NCGM, the number of COVID-19 patients during the fifth wave (n=22, 2 to 5 months after vaccination program in the hospital) was slightly higher than that during the third wave (n=17, before the vaccination program) despite 3.5 times the number of cases recorded during the former period than the latter period in Tokyo as a whole.^15^ The vaccine effectiveness against the Delta variant infection after 4 months was 53% in the U.S.^4^ and 35.1% in Qatar.^5^ It is clear that a hospital-wide vaccination program alone cannot eliminate the risk of the infection by the Delta variant. Still, we could reasonably infer that the program has contributed to the sizable reduction in the number of COVID-19 patients among the staff during the largest epidemic.

We confirmed that the breakthrough cases occurred independently in different timings and departments, showing no evidence of clustered infection in the hospital. This finding is consistent with our previous reports from the first and second serological surveys in NCGM before the vaccination program,^12,13^ suggesting that infection among the staff had occurred mainly outside the hospital (household, community, etc.). NCGM has adopted comprehensive measures against nosocomial infection since the early phase of the epidemic. The current data confirm the significant role of these measures to protect healthcare workers against the Delta variant infection.

Contrary to our prior expectation, we observed no measurable difference in the level of both neutralizing and anti-spike antibodies between cases with breakthrough infection and their controls. This finding contradicts with data from Israel^7^ and Vietnam,^8^ which reported significantly lower neutralizing antibody titers among those with breakthrough infection than controls. While the reason for the discrepant findings is not clear, it could be ascribed to the difference in the timing of blood sampling for antibody testing (peri-infection^7^ or pre-infection [present study]) the predominant type of variants (Alpha^7^ and Delta^8^), measurement of neutralizing antibody (use of three predominant types of live virus [present study] or use of unspecific surrogate-virus^7,8^], and case-control matching strategy.

Neutralizing capacity against the Alpha and Delta variants in the serum of the vaccinated participants was much lower than that against the wild-type strain, irrespective of breakthrough infection. This finding agrees with previously report^16^ and is consistent with the lower vaccine effect observed during the epidemic predominantly of the Variants of Concern.^3-5^ More than 90% of NCGM-Toyama ward staff have completed the second dose of vaccine by mid-April 2021. Soon after, the fourth epidemic wave of COVID-19 attacked Japan, dominated by Alpha; however, no vaccinated staff suffered from COVID-19, confirming high vaccine effect against infection of the Alpha variant within 2 months of vaccination.^4,5^ Breakthrough infection in NCGM from July to August 2021 could be attributed to the subsequent waning of neutralizing capacity and the largest epidemic due to the highly vaccine-resistant Delta variant.^9,16^

We found that all the COVID-19 patients who completed two-dose vaccination experienced no or only mild symptoms and had successfully returned to work, a finding compatible with the previous reports.^5,7,8,16^ These data support that vaccination contributes to the maintenance of hospital function by shortening sick leave due to COVID-19 of healthcare workers and facilitating their earlier return to work.

The present study has several strengths. Both cases and controls were derived from a well-defined cohort. We rigorously matched each case and control using the propensity score estimated by several factors potentially associated with infection risk (including the occupational risk of infection and prevention practices). The blood samples for antibody assays were obtained before the infection. We measured neutralizing antibody titers against the wild-type strain and two major variants using live viruses.

We also acknowledge study limitations. We measured antibody levels using samples obtained 5 to 10 weeks (median, 8 weeks) before the infection, which might have decreased at the time of infection due to the waning of antibodies over time. Nevertheless, we confirmed no evidence of the difference between cases and controls in the association of the duration of time between vaccination and blood sampling with neutralizing antibody titers (**Supplementary Figure 1**), denying faster waning of antibodies among cases than among controls. Data on virus type were available for only 5 cases (all Delta variants). However, the remaining breakthrough infections were most likely due to the Delta variant, which accounted for more than 90% of sequenced COVID-19 samples in Japan during the fifth epidemic wave.^10^

### Conclusion

In conclusion, pre-infection neutralizing antibodies against the wild-type, Alpha, and Delta variants did not materially differ between healthcare workers who experienced breakthrough infection and those who did not under the Delta variant rampage. The result points to the importance of continued adherence to infection control measures in the post-vaccination variant-circulating era, irrespective of the level of immunogenicity.

## Data Availability

The data is not publicly available due to ethical restrictions and participant confidentiality concerns, but de-identified data can be available for interested researchers after permission for using the data from the National Center for Global Health and Medicine Ethics Committee.

## Funding

This work was supported by the NCGM COVID-19 Gift Fund (grant number 19K059) and the Japan Health Research Promotion Bureau Research Fund (grant number 2020-B-09).

## Acknowledgments

We thank Haruka Osawa, Mika Shichishima, Yumiko Kito, and Azusa Kamikawa for their contribution to data collection and the staff of the Laboratory Testing Department for their contribution to measuring antibody testing.

## Declaration of Competing Interest

Abbott Japan and Roche Diagnostics provided reagents for anti-spike antibody assays.

## Notes

### Author Declarations

The ethics committee/IRB of the National Center for Global Health and Medicine gave ethical approval for this work.

