## Supplementary Appendix for "COVID-19 breakthrough infections and pre-infection neutralizing antibody"

### **Supplemental Text 1.** Algorism of case-control / propensity score matching

The controls were selected from the serological survey participants, who had received two doses of the vaccination and had data on the questionnaire and serological testing results (n=2,456). We excluded participants from control selection according to the following rules: those who had a history of COVID-19 diagnosis (n=14); Seropositive on IgG N protein by the serological survey (n=40). Thus, leaving 2,415 were included as the control source cohort.

For each breakthrough case, we matched three samples from the control source cohort based on the worksite, sex, the interval between the 2^nd^ vaccination and blood sampling, age (± three years), and propensity score (three closest absolute caliper values).

As the interval of the cases increase, greater differences among their controls are allowed with the following matching caliper. For cases with a time interval of up to 20 days, ±3 day difference is allowed for matched controls, for 21–40 days, ±5 days is allowed, for 40–80 days, ±10 days are allowed and for longer intervals (>80 days), up to ±15 days is allowed.

We estimated propensity score with the use of a multivariable logistic regression model, the presence/absence of breakthrough infection as the dependent variable, and the following variables as covariates: body mass index (continuous), occupational exposure risk of SARS-CoV-2 (low, moderate, high), use of public transportation (<1, 1-2, 3-4, ≥5 times/week), frequency of experiences spending more than 30 minutes in the 3Cs (crowded places, close-contact settings, and confined and enclosed spaces) without masking in leisure time (none, 1-2, 3-5, 6-9, ≥10 times), frequency of experiences having dinner with ≥5 people and ≥1 hour (none, 1-2, 3-5, 6-9, ≥10 times), and the adherence to 6 types of infection preventive behaviors (always, often, seldom, not at all). Infection preventive behaviors including (1) avoiding 3Cs; (2) social distancing (2 meters; if not possible, 1 meter); (3) wearing a mask when talking or when you are outdoors; (4) practicing good cough etiquette; (5) trying not to touch eyes, nose, and mouth; and (6) washing or sanitizing hands.

### **Supplemental Text 2.** The sources of cells and virus strains

VeroE6_TMPRSS2_ cells were obtained from Japanese Collection of Research Bioresources (JCRB) Cell Bank (Osaka, Japan) and maintained in Dulbecco’s Modified Eagle Medium (DMEM) supplemented with 10% FCS, 100 µg/ml of penicillin, 100 µg/ml of streptomycin, and 1 mg/mL of G418. As the wild-type strain, SARS-CoV-2 NCGM-05-2N strain (SARS-CoV-2_05-2N_) was isolated from nasopharyngeal swabs of a patient with COVID-19, who was admitted to the NCGM hospital in the early phase of the COVID-19 epidemic in 2020. Two clinically isolated SARS-CoV-2 mutant strains were used in the current study: the B.1.1.7 (Alpha) strain [hCoV-19/Japan/QHN001/2020 (SARS-CoV-2_QHN001_, GISAID Accession ID; EPI_ISL_804007)] was obtained from the National Institute of Infectious Diseases, Tokyo, Japan.; and the B.1.617.2 (Delta) strain [hCoV-19/Japan/TKYK01734/2021 (SARS-CoV-2_1734_, GISAID Accession ID; EPI_ISL_2080609)] was provided from Tokyo Metropolitan Institute of Public Health, Tokyo, Japan. Each variant was confirmed to contain each VOC-specific amino acid substitution before the assays conducted in the present study.

### **Supplemental Table 1.** Detailed information on each case of breakthrough infection in the case-control study

| **Sex** | **Age** | **Date of diagnosis** | **Reasons for diagnosis** | **Details of symptom** | **Virus strain** | **Ct value** | **Close contact** | **Source of infection** | **Treatment place** | **ICU admission** | **Use of ventilator/ecmo** | **Return to work** |
| --- | --- | --- | --- | --- | --- | --- | --- | --- | --- | --- | --- | --- |
| Men | 20s | July-21 | Symptoms | fever | Delta | - | Yes | community | Home | None | None | reinstatement |
| Men | 30s | July-21 | No symptom (close contact) | fever, cough, nasal discharge, malaise | Unknown | - | Yes | household | Home | None | None | reinstatement |
| Men | 20s | August-21 | Symptoms | fever, cough | Delta | - | Unknown |  | Home | None | None | reinstatement |
| Men | 20s | August-21 | Symptoms | fever | Delta | 16.23 | Unknown |  | Home | None | None | reinstatement |
| Men | 20s | August-21 | Symptoms | fever | Unknown | - | Unknown |  | Home | None | None | reinstatement |
| Women | 20s | August-21 | Symptoms | fever | Delta | - | Unknown |  | Home | None | None | reinstatement |
| Women | 20s | August-21 | Symptoms | sore throat, malaise | Delta | 24.1 | Unknown |  | Home | None | None | reinstatement |
| Women | 20s | August-21 | Symptoms | sore throat | Unknown | 37.8 | Yes | household | Home | None | None | reinstatement |
| Women | 20s | August-21 | Symptoms | fever, cough | Unknown | - | Yes | community | Home | None | None | reinstatement |
| Men | 40s | August-21 | Symptoms | nasal discharge, sore throat | Unknown | 23.56 | Unknown |  | Home | None | None | reinstatement |
| Women | 40s | August-21 | Symptoms | fever | Unknown | - | Yes | community | Home | None | None | reinstatement |
| Women | 40s | August-21 | Symptoms (close contact) | sore throat | Unknown | 16.13 | Yes | household | Home | None | None | reinstatement |
| Women | 40s | August-21 | Symptoms (close contact) | sore throat, cough | Unknown | 29.21 | Yes | household | Home | None | None | reinstatement |
| Women | 40s | August-21 | Symptoms (close contact) | fever, sore throat, nasal discharge | Unknown | 12.03 | Yes | household | Home | None | None | reinstatement |
| Women | 40s | August-21 | No symptom (close contact) | No symptom | Unknown | - | Yes | household | Home | None | None | reinstatement |
| Men | 20s | September-21 | Symptoms | fever | Unknown | 17.7 | Yes | community | Home | None | None | reinstatement |
| Men | 40s | September-21 | Symptoms | fever, malaise | Unknown | 11.19 | Unknown |  | Home | None | None | reinstatement |

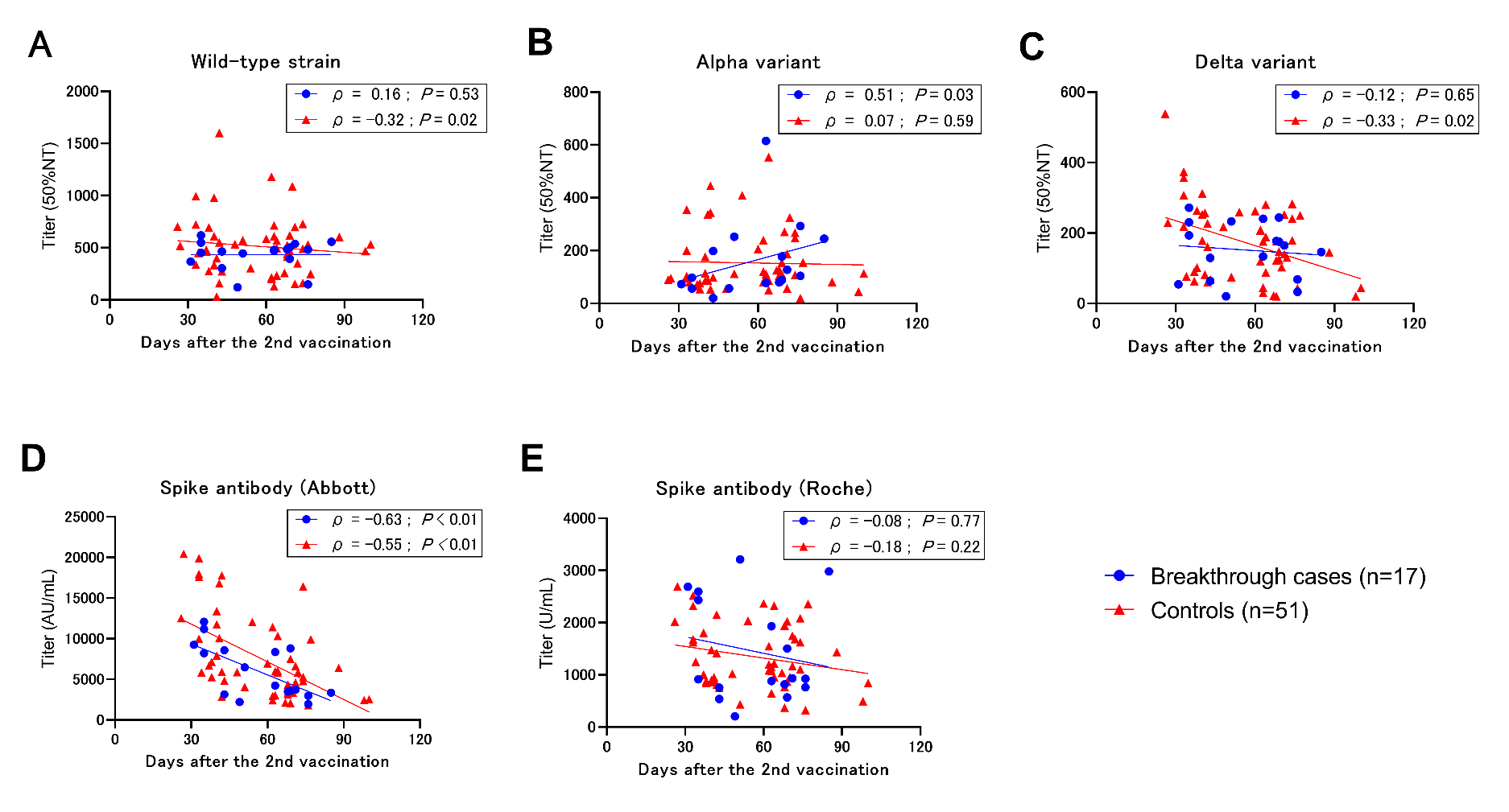

### **Supplemental Figure 1.** Correlations of neutralizing and anti-spike antibody titers with the timing of blood sampling after the second vaccination among breakthrough cases (n=17) and controls (n=51).

The Y-axis indicates 50% neutralizing titers against the wild-type strain (Panel A), the Alpha (B.1.1.7) variant (Panel B), or the Delta (B.1.617.2) variant (Panel C), or anti-spike antibody titers measured by Abbott (Panel D) and Roche (Panel E).

The X-axis represents the days between the second vaccination and the blood collection for serological testing of each subject.

ρ: Spearman’s correlation coefficient.
